# Morphological Patterns Of Anemia Among Under Five Children On Prevention Of Mother-To-Child Transmission (PMTCT) Programmes In Masogo Sub-County Hospital, Kisumu County, Kenya

**DOI:** 10.1101/2021.02.04.21250383

**Authors:** Silas O. Awuor, Omwenga O. Eric, Stanslaus Musyoki, Ibrahim I. Daud, Robert O. Nyangaresi, Peter Mugah, Beatrice Mukunzi

## Abstract

**Background:** Anaemia during childhood adversely affects mental, physical and social development of the children, therefore morphological patterns of anaemia in under-five children are considered essential for classification, diagnosis and management.

**Aim:** This study aimed at assessing morphological patterns, the prevalence and associated factors of anaemia among under-five children on Prevention of Mother-To-Child Transmission (PMTCT) programmes in Masogo sub-county hospital, Kisumu County, Kenya.

**Method:** A cross-sectional health facility-based study was conducted among 175 children aged 6 to 59 months who attended clinic for the PMTCT programme for the period of January 2020 to December 2020. Pretested and structured questionnaires were used to collect socioeconomic and demographic characteristics of the family and child. Capillary blood sample was collected from each child for malaria parasite and Peripheral Blood Film (PBF) examination.

**Result:** Complete blood counts indicate that microcytic pattern was the most common, representing 30 (42.3%) followed by microcytic hypochromic pattern 20 (28.2%), normocytic normochromic pattern with 11 (15.5%) and lastly dimorphic pattern with 10 (14.0%). High prevalence of anaemia was observed in children who were urban dwellers (50.0%), in children whose mothers aged 18-27 years (44.0%) and had no formal education (48.1%). Besides, the high prevalence rate of anaemia was found among children with a family monthly income of less than 500 Ksh. (46.9%), early (<6 months) introduction of complementary foods (71.4%)

**Conclusion:** This study has revealed that the prevalence of anaemia in children less than five years is high and is a severe public health problem in the study area. Therefore, the policymakers should make a strategy that can reduce poverty and increase the awareness to women on breastfeeding, nutrition, and other associated factors to reduce anaemia.

## 1.0 Introduction

Anaemia can be defined as a reduction in hemoglobin (Hb) concentration, hematocrit, or a number of red blood cells per litre below the reference interval for healthy individuals of similar age, sex, and race, under similar environmental conditions [1]. According to the World Health Organization (WHO), for under-five children, the threshold Hb level for being anemic is less than 110 g/l [2]. Anaemia is a global public health problem which affects 1.62 billion (24.8%) people world-wide. It occurs at all stages of the life cycle but is more prevalent in under-five years old children. Globally, 293.1 million (47.4%) under-five year’s children are anemic and 67.6% of these children live in Africa [3, 4]. In Kenya, 57% of children age 6–59 months were documented to be anemic according to the Kenya 2019 Demographic and Health Survey (KDHS) report [5]. This therefore raises a pertinent concern that calls for proper measures to be put in place to reduce such cases.

Several factors have been found to be contributing to the occurrence of anaemia and nearly half of (43%) the anaemia cases in childhood are due to iron deficiency [6]. The deficiency may result from inadequate dietary intake of iron, malabsorption of iron, and increased iron demand during rapid growth in children and chronic blood loss. Other causes of anaemia include folate and vitamin B12 and A deficiencies, Malaria, intestinal helminths, viral infections, chronic disease, hemoglobinopathies, hemolysis, and bone marrow disorders [7–10]. For instance, *H. pylori* has been documented to absorption of iron and its hypothesize that gastritis and disturbance in pH increase levels of neutrophil-derived lactoferrin, and since *Helicobacter pylori* has a lactoferrin-binding protein receptor, the infection would result in increased iron losses related to bacterial turnover [39]. Different studies also claimed that factors such as age, sex, residence, early initiation of complimentary food, under-nutrition, maternal health status, maternal education, and poor socioeconomic status are significantly associated with anaemia [11–13].

Anaemia on childhood adversely affects mental, physical, and social development of the children in short- and long-term outcome; it causes abnormalities of immune function, poor motor and cognitive development, poor school performance, and reduced work productivity in the life of the children, thereby decreasing earning potentials and negatively affect national economic growth. [14–17]. Anaemia is also an important cause of morbidity and mortality in African children where resources to determine the underlying etiology remain poor [18]. Based on the morphology of RBCs and blood cell indices, anaemia is classified into normocytic normochromic, microcytic hypochromic, macrocytic and dimorphic anaemia. Each type suggests specific aetiological factors, so an evaluation of the morphology of RBCs and clinical features among under five could help in the diagnosis and management of patients. Even though the national and regional prevalence of anaemia in under five years children are available in Kenya, data on the magnitude of anaemia and its risk factors in specific settings are scarce. Studying the specific aetiology and prevalence of anaemia in each setting and population group is very important to prevent or treat anaemia [2]. Therefore, this study is aimed to assess the morphological patterns of anaemia and its associated factors among children under-five years of age in Masogo sub-county hospital, Kisumu County, Kenya.

## 2.0: Materials and Methods

### 2.1 Study site

The study was conducted at Masogo Sub-County Hospital (0.1566^0^ S, 35.1984^0^ E) in Kisumu County (East to Chemelil Sugar Company). Masogo Sub-County Hospital is a Government a 20bed capacity health facility located in Masogo centre, Muhoroni Sub County in Kisumu County.

### 2.2: Study design

This was a cross-sectional study conducted at Masogo sub-county hospital, Kisumu County from January 2020 to December 2020. One hundred and seventy-five women with under five children attending PMTC clinics in the hospital were included, after fulfilling specific selection and exclusion criteria, in which only those mother who are living with HIV having under-five children and having ccc number with the facility will be consider in the study.

### 2.3: Sample size

### 2.4: Data collection on socio demographics

Pretested and structured questionnaires were used to collect socioeconomic and demographic characteristics of the family and child, feeding practice and other risk factors by interviewing mother/caregivers of the child. The questionnaire was adapted from previous similar literatures [12].

The questionnaire was pretested in Nyakoko health centre that was not included in the actual study area on 10% of the sample size. Based on the pilot study result, certain revisions were made for the questionnaire before the actual study. The interview was conducted by two trained clinical nurses in the local language (Dholuo).

### 2.5: Determination of the nutritional status

Data on nutritional status were collected by measuring the weight and height of children below age 5 during the clinic visit based on WHO recommendations [19]. The length was measured for children aged 6–23 months in a recumbent position and standing height was measured for children aged 24–59 months using the measuring board. The weight of the children was measured by a Salter scale as used before [19]. Briefly, the children were requested to remove shoes and any other heavy clothing prior to their weight measurement. Each measurement was collected twice and the mean value of the two measurements was recorded on the questionnaire.

### 2.6: Laboratory Analysis

#### 2.6.1: Venous blood collection

Venous blood was collected by use of well-established standard protocols as used before [20] by a well-trained phlebotomist. The vein was identify and the site was clean using 70% Alcohol, by used of 10ml syringe nine mL of venous blood was collected and divided into three containers: ethylene diamine tetra acetic acid (EDTA), lithium heparin and Plain containers.

#### 2.6.2: Determination of presence of anaemic blood parameters

##### i. Complete blood counts

The complete blood count was measured using Sysmex KX20 automated analyser, manufacturer by Sysmex Corporation, September 2015 (Bellport, New York, United States), and direct peripheral smears were taken to be compared with Ethylene diamine tetra-acetic acid (EDTA) smears; both thin and thick smears were stained with Gemsa stain and examined under the microscope for RBC morphological classification using already established standard protocols [20].

##### ii. Haemoglobin concentration determination

Two ml of blood in the plain tube were used to determine haemoglobin concentration using HemoCue Hb 201 analyzer as used before (Insert references). This analyzer uses HemoCue cuvettes Hb-201+ cuvettes which contains sodium deoxycholate dried reagent that lyses red blood cells to release free Hb and form a stable azidemethemoglobin that is detected at 570 nm and 880 nm. One drop of blood was carefully collected in a microcuvette from the sample on the plain container. The filled microcuvette was loaded in the cuvette holder of calibrated HemoCue Hb201analyser and after few seconds the haemoglobin measurement displayed. Then the results were recorded on the questionnaire. Standard operating procedure and manufacturers instruction were strictly followed [20, 21].

### 2.7: Data analysis procedures

All collected data were entered into Ms Excel and exported to SPSS version 20.0 statistical software for analysis. Normally distributed and continuous variables expressed as mean ± SD, and non-normally distributed variables were presented as medians (quartiles 25 and 75%). Chisquare (×2) test was used to compare proportions. Multivariate logistic regression was used to calculate adjusted odds ratios (OR) and the corresponding 95% confidence intervals (CI). A pvalue < 0.05 was used to indicate statistical significance.

### 2.8: Ethical considerations

Confidentiality and privacy were strictly adhered to and no names of individuals were recorded or made known in the collection or reporting of information. The study was granted ethical clearance by Kisumu County ministry of health department and from the sub county MOH office (MSC/KSM/3421/20). The participants consented for taking part in the study and they consented to have the results of this research work published.

## 3.0: Result

### 3.1: Sociodemographic characteristics

A total of 175 under-five children participated in the study from January 2020 to December 2020.The mean age of the children who participated in the study was 23.1±14.4 months (median: 20 months, range: 6 to 58 months). Half of the children were below two years of age 97 (55.4%), 127 (72.5%) of the children came from the rural part of the study area. Majority 91 (52.0%) of caregivers/mothers of the children were aged between 18 – 27 years old, followed by 81 (46.3%) age 28-38 years and lastly 3 (1.7%) age39-40 years. On education 104(57.4) had no formal education followed with 47(26.9%) with primary level and 24 (13.7%) had secondary and above. On occupation 132(75.5%) were Housewives with mean (±SD) mothers/caregivers age of 27.1 (± 5.3) years. Majority 131 (74.7%) of the mothers had one child aged under five years, and 136 (77.7%) of the studied children had first birth order. The median (interquartile range) monthly income of the families of the studied children was 800 (600–1050) Kenya shillings (Ksh), and 101 (57.7%) had a monthly income between 750 and 1500 ETB (Table 1).

**Table 1:**
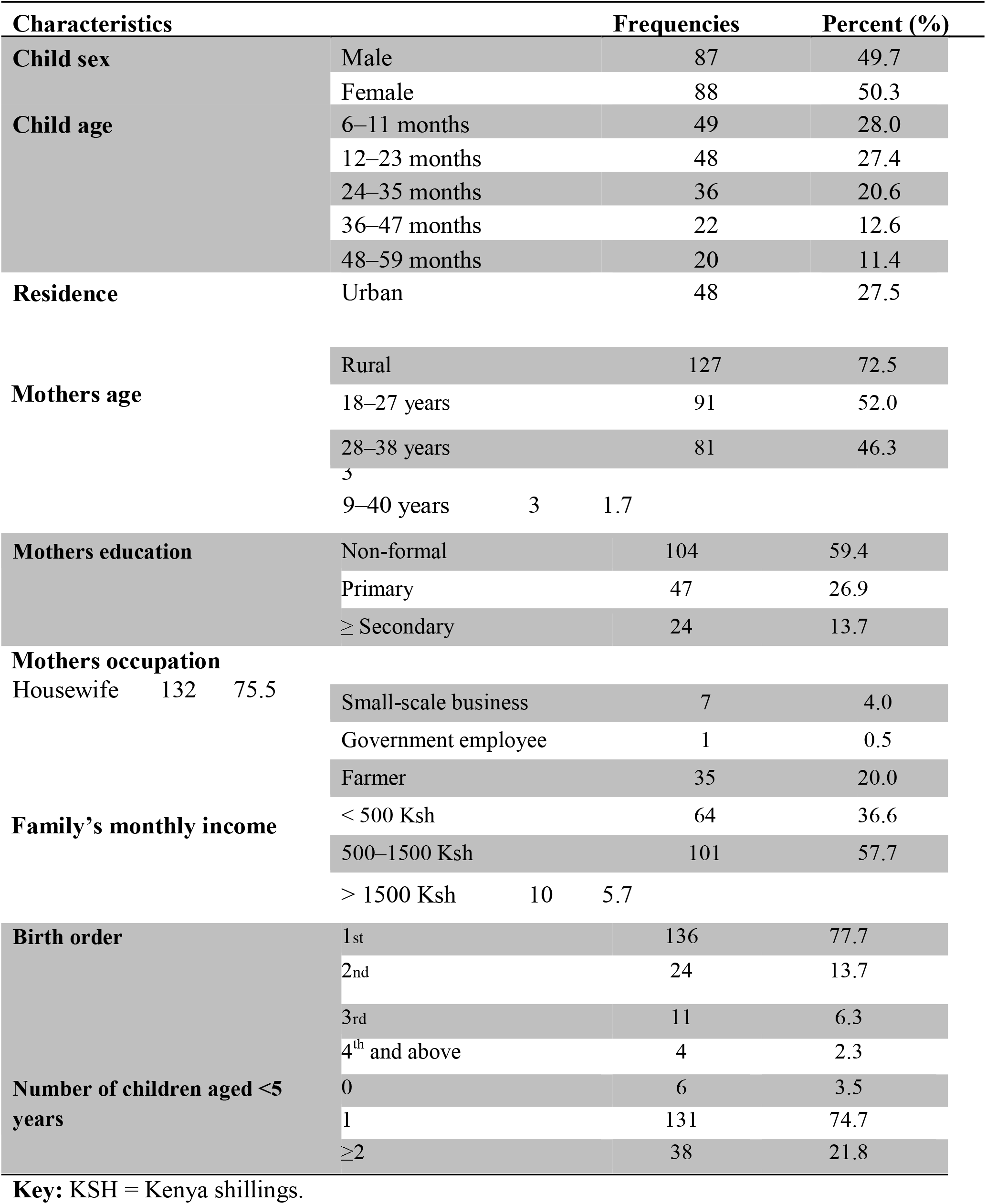
Socio-demographic characteristics of children under five years and mothers attending Masogo sub-county Hospital PMTC, (*n* = 175).

### 3.2: Nutritional status of children

Regarding nutritional status, 93(53.1%) of the children were underweight, 63(36%) were wasted in which 27 (15.4%) were moderate while 36 (20.1%) were sever, and 93(53%) were stunted, where 34 (19.4%) were moderate while 59 (33.3%) were sever. Only 14 (7.8%) of the children had started complementary foods before 6 months of their age. The blood sample for malaria parasite investigations showed that 72 (41.1%) of children suffered from malaria infestations (Table 2).

**Table 2:**
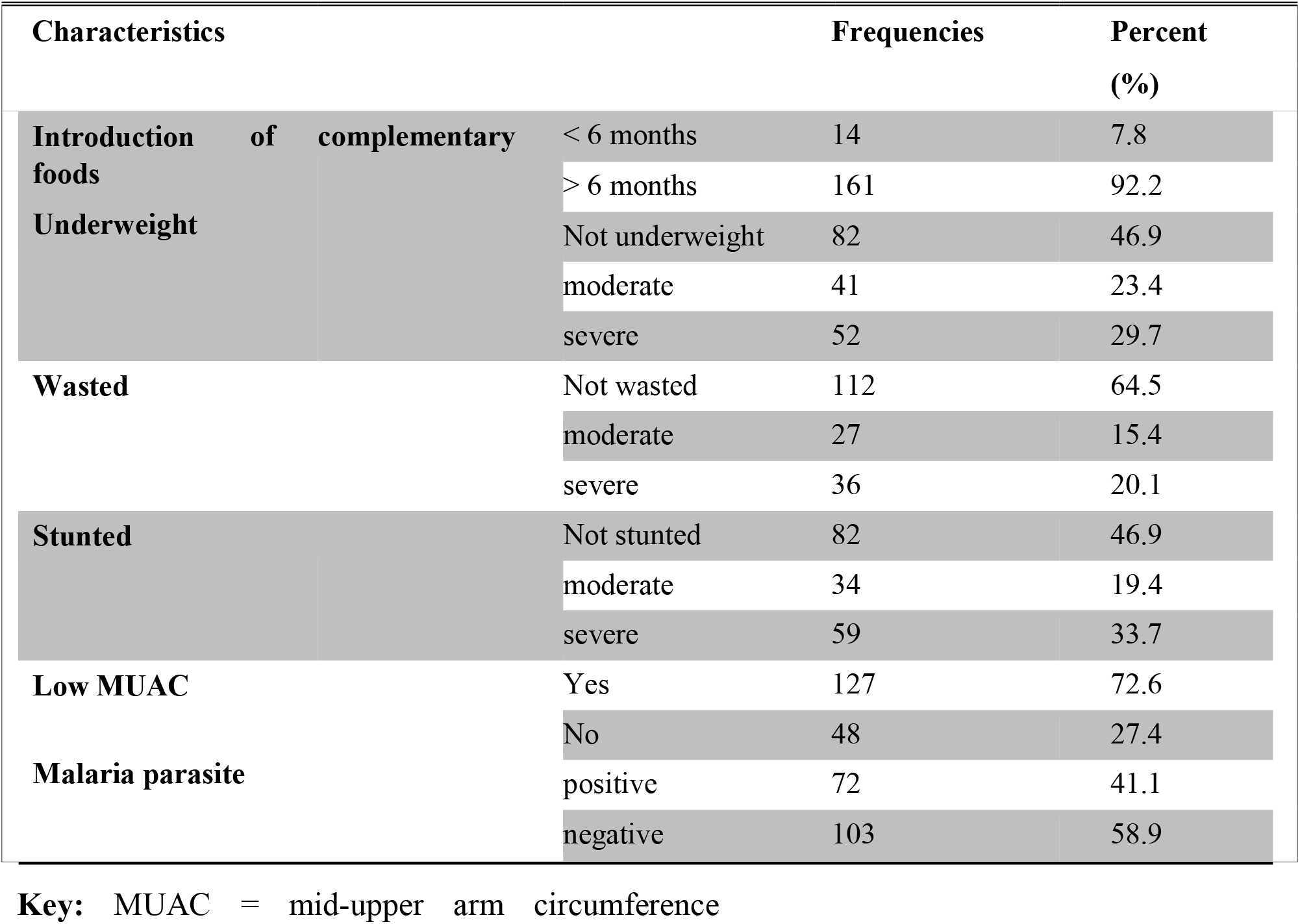
Anthropometric status and Malaria parasite infection of children under five years attending at Masogo sub-county hospital, (n = 175).

| Characteristics |  | Frequencies | Percent (%) |
| --- | --- | --- | --- |
| Introduction of complementary foods | < 6 months | 14 | 7.8 |
|  | > 6 months | 161 | 92.2 |
| Underweight | Not underweight | 82 | 46.9 |
|  | moderate | 41 | 23.4 |
|  | severe | 52 | 29.7 |
| Wasted | Not wasted | 112 | 64.5 |
|  | moderate | 27 | 15.4 |
|  | severe | 36 | 20.1 |
| Stunted | Not stunted | 82 | 46.9 |
|  | moderate | 34 | 19.4 |
|  | severe | 59 | 33.7 |
| Low MUAC | Yes | 127 | 72.6 |
|  | No | 48 | 27.4 |
| Malaria parasite | positive | 72 | 41.1 |
|  | negative | 103 | 58.9 |

### 3.3: Anaemia parameters

#### i. Complete blood counts

Complete blood counts did indicate that microcytic pattern was the most common, representing 30 (42.3%) followed by microcytic hypochromic pattern 20 (28.2%), normocytic normochromic pattern with 11 (15.5%) and lastly dimorphic pattern with 10 (14.0%) as shown in plates below (Fig.1). The general distribution of the blood cells is also summarized in Figure 2 below

**Figure 1:**
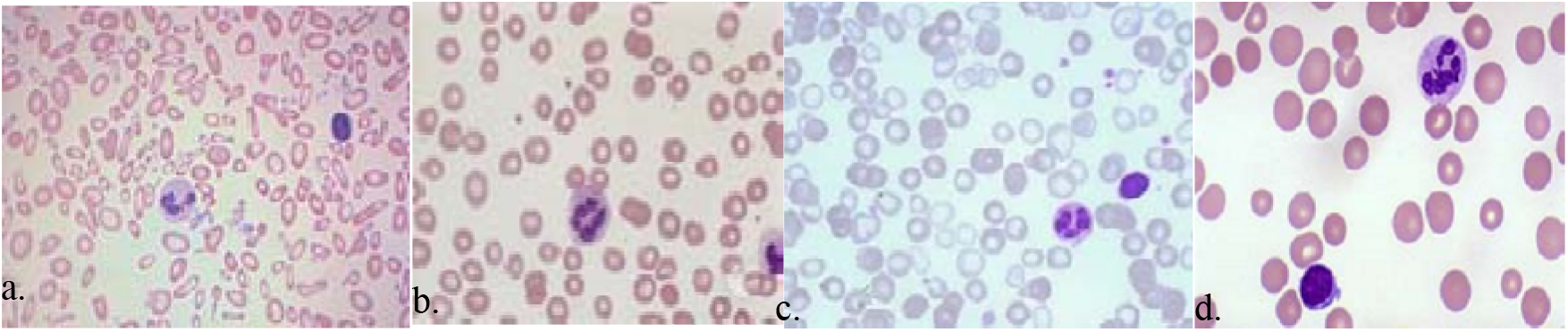
Photos of morphological patterns of anaemia, Masogo sub-county hospital, January 2020 to December 2020. (Plate a) Hypochromic microcytic, (Plate b) normochromic normocytic, (Plate c) dimorphic pattern and (Plate d). Macrocytic pattern

**Figure 2:**
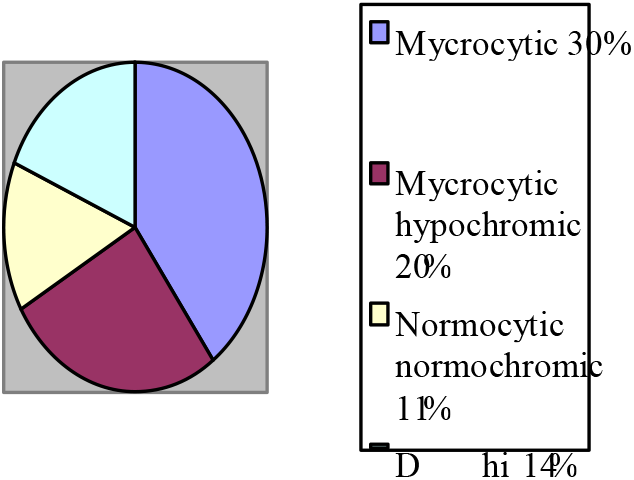
The general distribution of morphological patterns of anaemia, Masogo sub-county hospital, January 2020 to December 2020.

#### ii. Haemoglobin concentration

Haemoglobin concentration below 5 g/dl was high among the female 37 (19.5%) and male of 34 (19.5%), whereas the concentration of 5-10g/dl female were 35 (20%) and male were 23 (13.1%) and lastly the concentration of >10 g/dl male were 30 (17.1%) and female were 16 (9.1%) as summarize in table 3.

**Table 3:** Haemoglobin concentration among children under five years attending Masogo subcounty hospital.

| Characteristics |  | Frequency (n=175 | Percentage (%) |
| --- | --- | --- | --- |
| <5 g/dl | Male | 34 | 19.5 |
|  | Female | 37 | 21.2 |
| 5-10 g/dl | Male | 23 | 13.1 |
|  | Female | 35 | 20.0 |
| >10 g/dl | Male | 30 | 17.1 |
|  | Female | 16 | 9.1 |

### 3.4: Anaemia prevalence

High prevalence of anaemia was observed in children who were urban dwellers (50.0%), in children whose mothers aged 18-27 years (44.0%) and had no formal education (48.1%). In addition, the high prevalence rate of anaemia was found among children with a family monthly income of less than 500 Ksh. (46.9%), early (<6 months) introduction of complementary foods (71.4%) (Table 4).

**Table 4:**
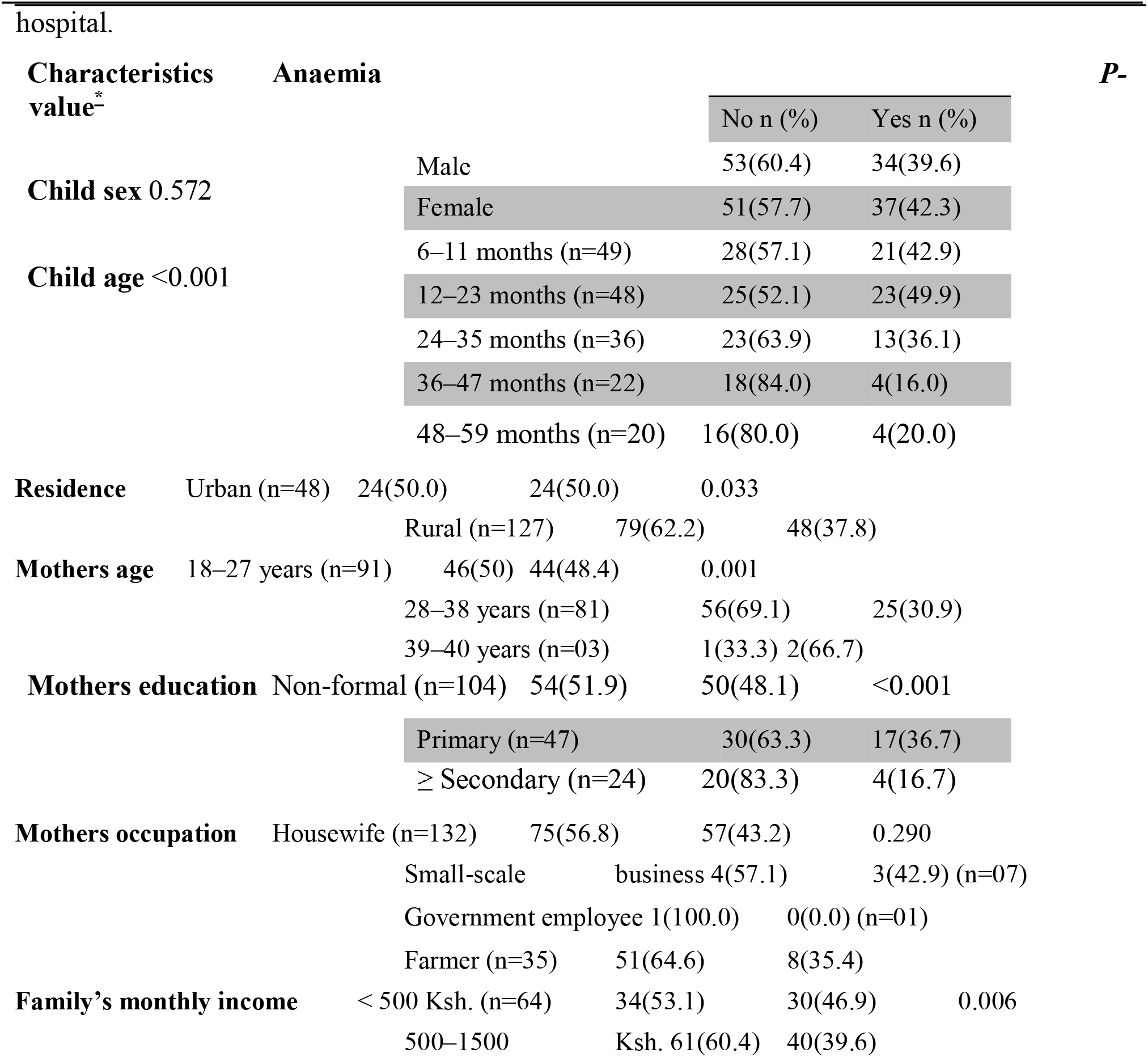
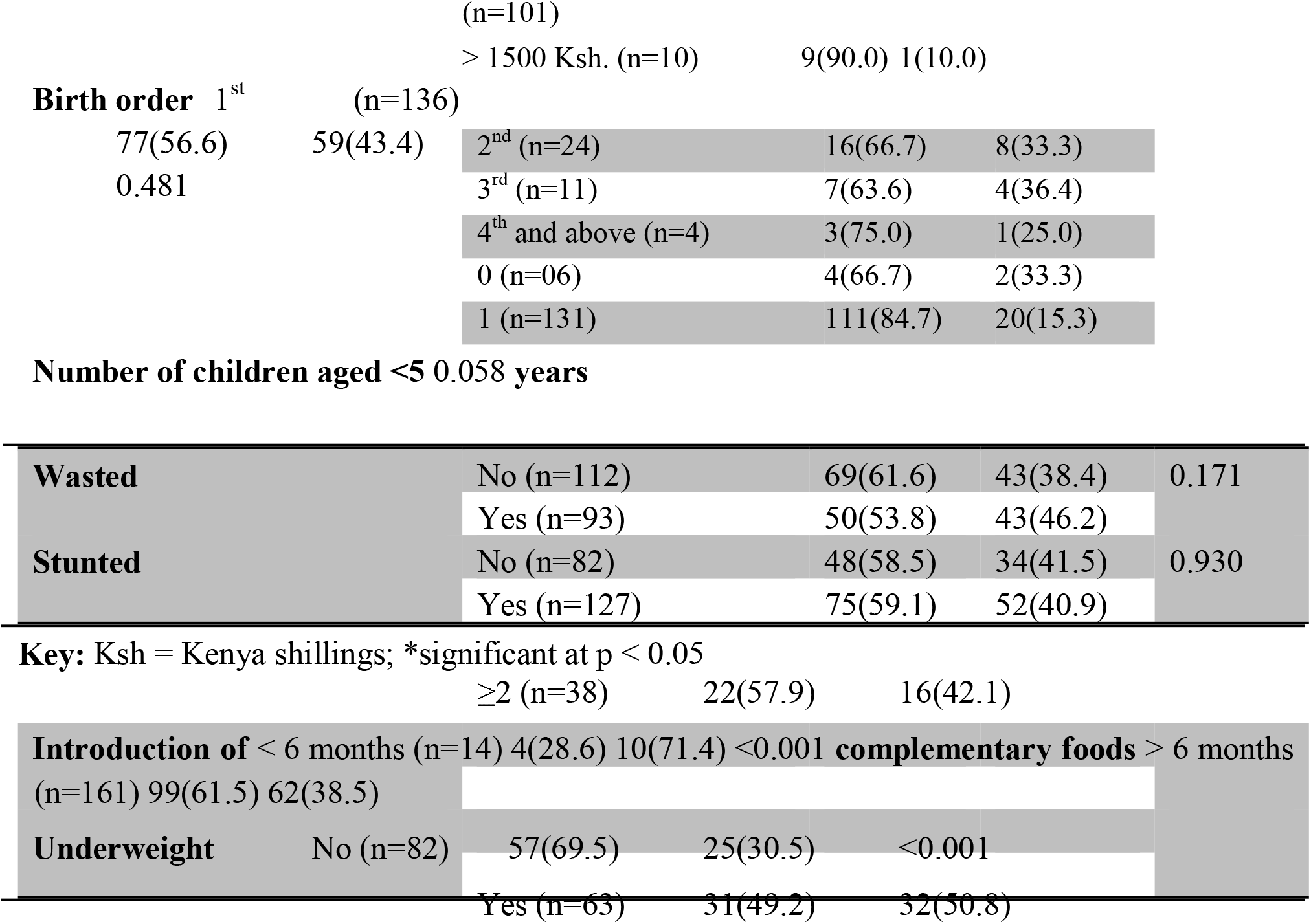
Prevalence of anaemia among children under five years attending Masogo sub-county

## Discussion

This study assessed the prevalence and associated factors of anaemia among children aged 6 to 59 months in Masogo sub county, Kisumu County. The overall prevalence of anaemia was 41.6%, a clear demonstration that it’s a common issue at the study area. This prevalence is higher as compared to that of WHO recommendations cut-off point of 11 g/dL in the same African children [41]. Our finding was similar to 2016 Ethiopian DHS prevalence reported for the Amhara Region (42%) [5] and study conducted in Jimma, Ethiopia (46%) [22]. However, the result of the present study is lower than studies conducted in Nepal (49.5%) [23], South-East Nigeria (49.2%) [24], Hohoe municipality and Volta Regional Hospital of Ghana (47.5% &55.0%) [25-26], and Limpopo Province, South Africa (75.0%) [27].The difference in the prevalence might be attributed in the study design, sampling techniques, and sample size. The difference might also be due to variation in the geographical location of the study participants or due to variation in socio-demographic characteristics or socioeconomic status of parents in the areas.

In this study, there was a higher prevalence of anaemia among children under two years old and it decreased as the age of the children increased. This could also be as a result of high iron demands associated with rapid growth rate and erythropoiesis, diets poor in bio-available iron, and low maternal iron reserve during pregnancy [33]. This finding is supported by other studies conducted in Ethiopia [28-30] and other developing countries [31-32] who also reported similar findings.

Like other study reports documented from South-East Nigeria [24] and the Volta Regional Hospital of Ghana [26], the present study found that sex difference did not show association with anaemia. However, other studies found a higher prevalence of anaemia among boys than girls [29], and also in girls than boys [27]. This inconsistency may be explained by the social norms in differential intake of iron-rich foods between genders; however, subsequent studies are required to better understand this complex issue.

Children of mothers with low educational levels were three times more likely to be anaemic than children of a mother with secondary and above education level. This may be explained by the fact that education enhances the mother’s knowledge needed for their children’s health and an appropriate feeding practice, which help to improve their children nutritional status [27]. However, this finding is inconsistent with the study conducted in Khartoum state, which showed the prevalence of wasting (3.3 to 21.1%) stunting (20.3 to 51.0%), severe stunting (12.9 to %) and underweight and severe underweight represent 24.4 to 35.0% and 6.6 to 48.0%, respectively [40]. [30]. The present study also found that children with low family income were highly more likely to be anaemic than those with high family income. A possible explanation for the high prevalence of anaemia might be that families with low income are less likely to purchase nutrient-rich foods (like iron, vitamins etc.), secure food availability, and afford health service during illness for their children. This finding was similar to studies conducted in other parts of the world, which reported that children from poor families were at risk of anaemia compared to their counterparts [11, 13, 30, 32, 34]. The higher prevalence of anaemia among children with less educated mothers and low-income families indicates that anaemia should be a marker of socioeconomic disadvantage. [36-37].

The observed higher prevalence of anaemia among children with early (<6 months) introduction of complementary foods was consistent with previous studies [11, 38]. Complementing breast milk before 6 months of age reduces the bioavailability of iron by up to 90%; the early introductions of complementary foods like cow’s milk interfering in the absorption of iron in the breast milk because cow’s milk has excess protein and minerals notably calcium and most digestive enzymes are inadequate at this age [11]. In addition, early initiation of complementary feeding exposes children to microbial pathogens due to contamination and resulting high risk of diarrheal diseases, thereby malabsorption [5]. Other pathogens like H. pylori has also been documented to possess a lactoferrin-binding protein receptor which has a higher affinity for Iron hence if present in the stomach can reduce the amount of iron being absorbed [39]. However, this finding is in contrary to study done in Sri Lanka which showed that children who were exclusively breastfed for 6 months or more were more likely to be anaemic than children who were exclusively breastfed for less than 6 months[35].

Nutritional status was also associated with anaemia among children aged 6–59 months. In this study, underweight children were most affected more likely to be anaemic than children with normal weight. This finding is similar to a study conducted in Northern Ethiopia [30] and Brazil [38]. Usually, the causes of anaemia and underweight (malnutrition) are similar and aggravated by poverty and food insecurity. Food insecurity affects the nutritional status of children by compromising the quantity and quality of dietary intake, which contributes for development of anaemia [6].

One of the limitations of this study is the cross-sectional nature of the study design; it does not reveal causal links between independent variables and anaemia. Due to constraint of resource, we were unable to measure serum ferritin concentration, soluble transferrin receptor concentration, folate levels, vitamin B12 levels, thalassemia, and G6PD deficiency, which could have helped in finding the causes of anaemia. The other limitation is that this study was conducted at a sub county hospital hence, further community based studies should be conducted to have findings more representing the whole population. Despite the limitations, we have determined the magnitude of anaemia and identified important factors associated with anaemia among children aged 6–59 months in Masogo sub county hospital.

## Data Availability

Any additional data required will be given

## Acknowledgement

We wish to sincerely acknowledge and thank MCH/PMTCT department members and laboratory department of Masogo sub county hospital for providing us the required data. We also thank Elsy jebitok for the assistance on the bench work on sample analysis and lastly the hospital in-charge for allowing us to do our research in the facility.

## Competing interests

The authors declare that no competing interests exist.

## Authors’ contributions

All authors contributed equally to this work.

## Funding information

This research received no specific grant from any funding agency in the public, commercial or not-for-profit sectors.

## Data availability statement

Data sharing is not applicable to this article as no new data were created or analysed in this study.

## Disclaimer

The views and opinions expressed in this article are those of the authors and do not necessarily reflect the official policy or position of any affiliated agency of the author

## Appendix

### Appendix I: Consent Form

My name is ***Silas Onyango Awuor*** I am a student at Kisii University pursuing Msc. In Medical microbiology. I am conducting a study on “**MORPHOLOGICAL PATTERNS OF ANAEMIA AMONG UNDER FIVE YEAR OLD CHILDREN ON PREVENTION OF MOTHER-TO-CHILD TRANSMISSION (PMTCT) PROGRAMMES IN MASOG SUB-COUNTY HOSPITAL, KISUMU COUNTY, KENYA**.” The information will be used by the Kisumu County Ministry of Health.

### Procedures to be followed

Participation in this study will require that you fill in a questionnaire. You have the right to refuse participation in this study. Please remember that participation in this study is voluntary. You may ask questions related to the study at any time. You may refuse to respond to any questions and you may stop an interview at any time.

### Risks

There are no risks whatsoever for participating in this study.

### Benefits

If you participate in this study you will help the Kisumu County Ministry of Health to know how to improve their health and therefore reduce the risks associated to anaemia.

### Confidentiality

You will fill the questionnaire on your own and your name will not be recorded on the questionnaire. The questionnaire will also be kept safe and everything will be kept private.

### Contact information

If you have any questions you may contact Silas Awuor 0723906204, Habakkuk Awino 729 60 3228.

### Participant’s Statement

The above information on my participation in the study is clear to me. I have been given a chance to ask questions and my questions have been answered to my satisfaction. My participation in this study will be entirely voluntary. I understand that my records will be kept private and that I can stop participation at any time.

Name of the participant ……………………………………………

__________

Signature Date

____________

Date

### Investigators Statement

I, the undersigned, have explained to the volunteer in a language s/he understands the procedures to be followed in the study and the benefits involved.

Name of the interviewer: ____________

____________

Interviewer

____________

Signature Date

## Notes

### Competing Interest Statement

The authors have declared no competing interest.

### Funding Statement

No funding for this work

### Author Declarations

Thestudy was granted ethical clearance by Kisumu County ministry of health department and from the sub county MOH office (MSC/KSM/3421/20)."

## References

1. Lanzkowsky P. Manual of pediatric hematology and oncology. 5th ed. Elsevier; 2005.

2. WHO. Haemoglobin concentrations for the diagnosis of anaemia and assessment of severity. Vitamin and Mineral Nutrition Information System. Geneva, World Health Organization, 2011. (WHO/NMH/NHD/MNM/11.1).

3. Benoist Bd, McLean E, Egll I, Cogswell M. Worldwide prevalence of anaemia 1993– 2005: WHO global database on anaemia. Geneva, World Health Organization. 2008.

4. McLean E, Cogswell M, Egli I, Wojdyla D, De Benoist B. Worldwide prevalence of anaemia, WHO vitamin and mineral nutrition information system, 1993–2005. Public health nutrition. 2009; 12(4):444–54.https://doi.org/10.1017/S1368980008002401 PMID: 18498676

5. Melku M, Addis Z, Alem M, Enawgaw B. Prevalence and predictors of maternal anaemia during pregnancy in Gondar, Northwest Ethiopia: An institutional based cross-sectional study. Anaemia. 2014;2014:1–9. https://doi.org/10.1155/2014/108593

6. Adish A, Esrey S, Gyorkos T, Johns T. Risk factors for iron deficiency anaemia in preschool children in northern Ethiopia. Public health nutrition. 1999; 2(3):243–52. PMID: 10512558

7. Lopez A, Cacoub P, Macdougall IC, Peyrin-Biroulet L. Iron deficiency anaemia. The Lancet. 2016; 387(10021):907–16.

8. Pacifici GM. Effects of Iron in Neonates and Young Infants: a Review. International Journal of Pediatrics. 2016; 4(7):2256–71.

9. Crawley J. Reducing the burden of anaemia in infants and young children in malariaendemic countries of Africa: from evidence to action. The American journal of tropical medicine and hygiene. 2004; 71 (2_suppl):25–34.

10. Janus J, Moerschel SK. Evaluation of Anaemia in Children. American Family Physician. 2010; 81(12):1462–1471. PMID: 20540485

11. Woldie H, Kebede Y, Tariku A. Factors associated with anaemia among children aged 6– 23 months attending growth monitoring at Tsitsika Health Center, Wag-Himra Zone, Northeast Ethiopia. Journal of nutrition and metabolism. 2015; 2015.

12. Ngesa O, Mwambi H. Prevalence and risk factors of anaemia among children aged between 6 months and 14 years in Kenya. PLoS One. 2014; 9(11):e113756. https://doi.org/10.1371/journal.pone.0113756 PMID: 25423084

13. Cardoso MA, Scopel KK, Muniz PT, Villamor E, Ferreira MU. Underlying factors associated with anaemia in Amazonian children: a population-based, cross-sectional study. PloS one. 2012; 7(5):e36341. https://doi.org/10.1371/journal.pone.0036341 PMID: 22574149

14. Grantham-McGregor S, Baker-Henningham H. Iron deficiency in childhood: causes and consequences for child development. Annales Nestle (English ed). 2010; 68(3):105–19.

15. Oppenheimer SJ. Iron and its relation to immunity and infectious disease. The Journal of nutrition. 2001; 131(2):616S–35S.

16. Lozoff B. Iron deficiency and child development. Food and nutrition bulletin. 2007; 28(4_suppl4):S560–S71.

17. Haas JD, Brownlie IVT. Iron deficiency and reduced work capacity: a critical review of the research to determine a causal relationship. The Journal of nutrition. 2001; 131(2):676S–90S.

18. Brabin BJ, Premji Z, Verhoeff F. An analysis of anaemia and child mortality. The Journal of nutrition. 2001; 131(2):636S–48S.

19. WHO. WHO child growth standards: length/height for age, weight-for-age, weight-forlength, weight for-height and body mass index-for-age, methods and development: World Health Organization; 2006.

20. National Committee for Clinical Laboratory Standards. H15-A3: Reference and selected procedures for the quantitative determination of hemoglobin in blood. 3. 2000.

21. HemoCue America. Hb-201+ Product Sheet. Available from: http://www.hemocue.us/~/media/hemocue-images/hemocue_usimages/pdf/hemoglobin-lit1056-hb-201product-sheet.pdf?la=enUS.

22. Getaneh T, Girma T, Belachew T, Teklemariam S. The utility of pallor detecting anaemia in under five years old children. Ethiopian medical journal. 2000; 38(2):77–84. PMID:11144886

23. Joshi S, Pradhan MP, Joshi U. Prevalence of anaemia among children under five years in tertiary care hospital of Nepal. Medical Journal of Shree Birendra Hospital. 2014; 13(1):33–6.

24. Ughasoro MD, Emodi I, Okafor H, Ibe B. Prevalence and risk factors of anaemia in paediatric patients in South-East Nigeria. South African Journal of Child Health. 2015; 9(1):14–7.

25. Parbey PA, Kyei-Duodu G, Takramah W, Tarkang E, Agboli E, Takase M, et al. Prevalence of Anaemia and Associated Risk Factors among Children Under Five Years in Hohoe Municipality, Ghana. Journal of Scientific Research & Reports. 2017; 15(2):1–

26. Adu-Amankwaah J, Allotey EA, Kwasie DA, Afeke I, Owiafe PK, Adiukwu PC, et al. Prevalence and Morphological Types of Anaemia among Children Under-Five Years in the Volta Regional Hospital of Ghana. Open Access Library Journal. 2018; 5(02):1.

27. Heckman J, Samie A, Bessong P, Ntsieni M, Hamandi H, Kohler M, et al. Anaemia among clinically well under-fives attending a community health centre in Venda, Limpopo Province. South African Medical Journal. 2010; 100(7):445–448. PMID: 20822592

28. Shakuntal G, Mane AS. Anaemia in pediatric patients under five years old: A crosssectional study. Scholars Journal of Applied Medical Sciences. 2016; 4(6):2020–2022.

29. Guled RA, Mamat NM, Balachew T, Bakar MA, Azdie W, Assefa N. Predictors and prevalence of anaemia, among children aged 6 to 59 months in shebelle zone, somali region, eastern Ethiopia: A cross sectional study. International Journal of Development Research. 2017; 7(1):11189–96.

30. Gebreegziabiher G, Etana B, Niggusie D. Determinants of anaemia among children aged 6–59 months living in Kilte Awulaelo Woreda, Northern Ethiopia. Anaemia. 2014; 2014:1–9.

31. Kuziga F, Adoke Y, Wanyenze RK. Prevalence and factors associated with anaemia among children aged 6 to 59 months in Namutumba district, Uganda: a cross-sectional study. BMC pediatrics. 2017; 17 (1):25–33. https://doi.org/10.1186/s12887-017-0782-3 PMID: 28100200

32. Khan JR, Awan N, Misu F. Determinants of anaemia among 6–59 months aged children in Bangladesh: evidence from nationally representative data. BMC pediatrics. 2016; 16(1):3–14.

33. Xin Q-Q, Chen B-W, Yin D-L, Xiao F, Li R-L, Yin T, et al. Prevalence of Anaemia and its Risk Factors among Children under 36 Months Old in China. Journal of tropical pediatrics. 2017; 63(1):36–42. https://doi.org/10.1093/tropej/fmw049 PMID: 27543970

34. Malkanthi R, Silva K, Jayasinghe-Mudalige UK. Risk factors associated with high prevalence of anaemia among children under 5 years of age in paddy-farming households in Sri Lanka. Food and Nutrition Bulletin. 2010; 31(4):475–82.

35. Alelign T, Degarege A, Erko B. Prevalence and factors associated with undernutrition and anaemia among school children in Durbete Town, northwest Ethiopia. Archives of Public Health. 2015; 73(1):34. https://doi.org/10.1186/s13690-015-0084-x PMID: 26261719

36. Allen L, de Benoist B, Dary O, Hurrell R. Guidelines on food fortification with micronutrients. Geneva: World Health Organization and Food and Agriculture Organization of the United Nations. 2006.

37. Ali D, Saha KK, Nguyen PH, Diressie MT, Ruel MT, Menon P, et al. Household Food Insecurity Is Associated with Higher Child Undernutrition in Bangladesh, Ethiopia, and Vietnam, but the Effect Is Not Mediated by Child Dietary Diversity. The Journal of nutrition. 2013; 143(12):2015–21. https://doi.org/10.3945/jn.113.175182 PMID: 24089419

38. Novaes TG, Gomes AT, Silveira KCd, Magalhães EIdS, Souza CL, Netto MP, et al. Prevalenceand factors associated with anaemia in children enrolled in daycare centers: A Hierarchical Analysis. Revista Paulista de Pediatria. 2017; 35(3):281–88. https://doi.org/10.1590/1984-0462/;2017;35;3;00008 PMID: 28977293

39. Nermine HZ., Ahmed EA. Investigation of a possible association between Refractory Iron Deficiency Anaemia to an Underlying Remote Helicobacter pylori Infection 2009. Journal of Egypt Public Health Association; 84:1–2.

40. Kanan SO, Abd Elmoneim OE. Prevalence and Causes of Undernutrition among Underfive Sudanese Children: A Mini-review. Journal of Advances in Medicine and Medical Research. 2020 Sep 3:12–20.

41. Kumar T, Taneja S, Sachdev HPS, Refsum H, Yajnik CS, Bhandari N, et al. Supplementation of vitamin B12 or folic acid on hemoglobin concentration in children 6– 36 months of age: A randomized placebo controlled trial. Clin Nutr. 2017 Aug;36(4):986–91. pmid:27486122

